# Translation, Adaptation, and Validation of Survey on Patient Safety Culture for Ambulatory Care in Japan

**DOI:** 10.64898/2026.02.05.26345699

**Authors:** Khin Zay Yar Myint, Ikumi Genka, Junichi Taguchi, Toshiomi Kusano

**Affiliations:** Tokyo Midtown Clinic, Minato-ku, Tokyo, Japan; Nihonbashi Muromachi Mitsui Tower Midtown Clinic, Chiyoda-ku, Tokyo, Japan

**Author notes:** Corresponding author and to whom reprint requests should be addressed Khin Zay Yar Myint, Tokyo Midtown Clinic, 9-7-1, Akasaka, Minato-ku, Tokyo, 107-6206.

**Keywords:** Safety culture, Ambulatory care, MOSPSC, Validation, Adaptation

## Abstract

**Objective:** There is no validated questionnaire in Japan to measure the culture of safety in ambulatory care clinics. Therefore, we developed the Japanese version of the Medical Office Survey on Patient Safety Culture (MOSPSC) of the Agency of Healthcare Research and Quality (AHRQ) in the United States with the aim to establish a tool for evaluating and benchmarking the safety culture of outpatient clinics in Japan.

**Materials and methods:** This research uses both qualitative and quantitative approaches to translate, adapt and validate the MOSPSC questionnaire which consists of 62 questions. The process involved seven steps such as translation by two independent bilingual physicians, drafting and reviewing, backtranslation by two separate translation companies, semantic equivalence assessment by AHRQ and revision, pretest, focused discussion, and finalizing the questionnaire after expert review and proofreading. An actual safety culture survey was conducted with mainly online and paper versions at four clinics in Tokyo. The survey results were then evaluated for patient safety dimensions, reliability and construct validity.

**Results:** Efforts are made to select appropriate terminology during tool adaptation processes due to different language and medical system between Japan and the United States. The response rate in the actual survey was 66.4% (242/364). Confirmatory factor analysis showed that factor loading and goodness of fit indices were better when 3 items were removed from the original 10-composite model with 38 items. The Cronbach’s alpha coefficients of composite measures ranged from 0.62 to 0.78 in the original model and 0.62 to 0.85 in the new model, indicating good internal consistencies.

**Conclusions:** Considering the differences in medical systems, culture, and language between the United States and Japan, the instrumented was adapted with a satisfactory content validity and reliability.

## Introduction

With increasing complexity and advances in medicine, modern healthcare has become vulnerable to errors. Consequently, fostering a culture of safety has been prioritized in medical institutions in recent years. Safety culture is a sub-facet of the organizational culture that pertains to safety and reflects an organization’s commitment to, and effectiveness in, health and safety management [1,2]. According to the Japanese Ministry of Health, Labour, and Welfare, safety culture in healthcare is defined as “A way of thinking or an attitude in which all the staff engaged in medical services give the highest priority to patient safety and strive for its realization, as well as the way organizations work toward that goal”[3].

Dr. James Reason identified five elements of safety culture: an informed culture, a reporting culture, a learning culture, a just culture, and a flexible culture [4]. The outcome of these safety cultures is reflected in medical errors, and it is important to analyze these errors from incident reports, mortality and morbidity conferences, and chart reviews [5–8]. As for a proactive approach to preventing errors, we can evaluate the pre-existing organizational culture in relation to safety, develop action plans for improvements, assess safety interventions and changes over time, and serve as internal and external benchmarkings [9–10]. However, measuring safety culture presents several challenges [11]. Measurements include surveys, document reviews, work observations, individual interviews, and focused groups [12]. Among them, surveys are practical and time-efficient and, most importantly, provide ease of benchmarking.

There are different safety culture surveys in healthcare [13]. The most widely adopted international patient safety culture surveys were developed by the Agency of Healthcare Research and Quality (AHRQ). The Japan Council for Quality Health Care has used the validated Japanese version of the Hospital Survey on Patient Safety Culture (HSOPS) by the AHRQ to evaluate the safety culture of hospitals in Japan [14, 15]. Conversely, there is no questionnaire in Japan to evaluate the safety culture of ambulatory medical facilities.

The AHRQ has a safety culture survey called “Medical Office Survey on Patient Safety Culture (MOSPSC)” for ambulatory care. As of September 2022, it has been translated into 15 languages and conducted in 38 countries [16]. Yet, there is no Japanese version of the MOSPSC. Moreover, it is critical to develop a reliable and valid instrument that can be implemented at ambulatory medical institutions in Japan to evaluate safety culture, implement effective measures according to the survey results, and provide benchmarking with other facilities. Therefore, we developed a Japanese version of the AHRQ’s MOSPSC to improve medical safety and quality in our group of clinics and expand its use for external benchmarking.

## Materials and methods

### Instrument

The MOSPSC consists of 62 questions, including free comments, and evaluates the safety culture of the respondents, the team they work for, their immediate supervisor, and the organization in sections A to H [17].

### Forward translation

We developed the Japanese version of the MOSPSC in seven steps: Step 1 was the translation of the original MOSPSC questionnaire into Japanese by two bilingual physicians whose native language is Japanese, one of whom knew the purpose of the translation, and the other who did not. In Step 2, these two translations were reviewed by a review committee consisting of the above two physicians, the principal investigator, and the chief nurse. The review committee identified noticeable differences in the classification and roles of medical professions between the United States and Japan and question items that were difficult to translate due to the use of double negatives. The review committee also referred to question items already included in the validated Japanese version of HSOPS for appropriate wording and structure [14]. After all these considerations, the two translations were modified and integrated into the first Japanese draft (T1).

### Back-translation and semantic equivalence

Step 3 was the back translation into English by two external translators from different translation companies who were native English speakers and did not know the original English version of the questionnaire. Step 4 involved examining the semantic equivalence of the two back-translated versions with the original English version. They pointed out five points and we responded accordingly. The details are described in Supplementary data file. Based on our responses, the AHRQ determined that the items closely followed the original survey, and we proceeded with the next steps in administering the survey at our medical facilities. In Step 5, based on the feedback from the AHRQ, the review committee modified the integrated version (T1) into a revised version (T2).

### Pilot test and focused discussion

In phase I of Step 6, we created T2 as a web questionnaire using the Microsoft form application. We then conducted a preliminary test with 34 employees at the Tokyo Midtown Clinic in Roppongi. The preliminary test included twelve medical staff, eight nurses, five administrative staff, three clinical technicians, three radiological technicians, two nurse aids, and one manager. In phase II of Step 6, a focused discussion with five staff members from five different professions (i.e., doctors, nurses, clinical and radiological technicians, and administrative staff) was conducted to evaluate the clarity of the questions and the selection of appropriate words. Details are provided in Supplementary data file. The questionnaire edited in accordance with the focused group discussion results was termed T3.

### Proofreading by an expert and actual survey

Finally, in step 7, an expert Japanese language proofreader assessed the grammar, style, clarity, pertinence, and understandability of the contents. He suggested changing all sentences from the polite form to the dictionary form in Japanese. We developed the final version of the questionnaire (T4) to conduct an actual survey at four clinics in the Tokyo area: Tokyo Midtown Roppongi, Nihonbashi, Ariake, and Tokyo Bay, all of which belong to the Medical Corporation Midtown Clinic. A survey link was provided to all staff working at these facilities. For those who did not have email addresses or personal computers at the workplace, paper survey questionnaires were distributed and patients were asked to drop them into a designated internal post box after completion. The survey was distributed and collected between September 21, 2023, to November 30, 2023. The timeline and flowchart for developing the questionnaire are outlined in Supplementary Figure file.

### Ethical considerations

The survey was conducted anonymously, in a self-administered manner. After explaining the purpose and other essential contents of the survey on the first page, we asked for consent. Only those who provided consent on the first page were included in the final analysis. The study was approved by the Institutional Review Board of The Midtown Clinic Medical Corporation (Rin-2021-10).

### Data analysis

First, we entered the data from the paper survey forms into a web form and saved the data file in Microsoft Excel. There was no missing data. We then coded the responses in accordance with the scales in each section: 1 to 5 in sections A and B, and 1 to 6 in sections C to G, with 9 coded for Does not Apply/Don’t know. The next step was to check the data file for possible data entry errors using frequency tables for each item to ensure that all responses were within a valid range. We followed the AHRQ’s MOSPSC data analysis guidelines to calculate the frequency percentages of positive responses to the single-item measures in sections A and B. We categorized the 38 items in sections C to F into 10 composite measures of patient safety culture and calculated the percent positive response, means, and composite measure scores. Negatively worded items were reverse-coded, as recommended by the AHRQ. The results were then compared with those from the AHRQ database.

Cronbach’s alpha coefficient and the average inter-item correlation (AIIC) were used to analyze reliability. We considered Cronbach’s alpha values >0.6 adequate for each composite, and at least 0.9 for the entire questionnaire. AIIC evaluates the correlation of items within each composite, and a value between 0.15 and 0.5 is considered evidence that the items are measuring the same underlying composite[18]. For structural validity, we performed confirmatory factor analysis (CFA) to determine how well the Japanese version fits the composite structure of the original MOSPSC. We did not perform exploratory factor analysis because we aimed to maintain the same factor structure as the original MOSPSC for benchmarking, and the MOSPSC has already been validated across different languages. First, we conducted Kaiser-Meyer-Olkin (KMO) and Bartlett tests of sphericity to evaluate whether our sample size was appropriate for factor analysis. The cut-off value for factor loadings was set to 0.40. We set the criteria for goodness-of-fit indices based on previous studies [19]. CFA was repeated if the factor loadings or model fit indices were not acceptable.

Finally, we calculated polychoric correlations for the 10 composites of patient safety culture to verify construct validity, as the items were ordinal variables. Figures and tables were generated using Microsoft Excel and PowerPoint. Comparison with AHRQ database was conducted using the Medical Office Survey Excel Tool Version 6.2 July 2024.20 Data analysis and statistical tests were performed using the R software (R version 4.4.2). The CFA was performed using the “*lavaan*” package, and the Cronbach’s alpha and AIIC calculations were performed using the “*psych*” package. Polychoric correlations and correlation plot were generated using the “*polycor*” and “*corrplot*” packages.

## Results

### Characteristics of participants

A total of 242 employees responded to the survey: 165 participated via the online form, and 77 returned the paper questionnaires. The response rate was 66.4% (242/364). We analyzed the data of the 222 participants who provided consent to participate in the study. Detailed characteristics of the participants are presented in Table 1. The rates of participation according to occupation were 61%, 27%, 71%, 74%, and 60% for physicians, leaders and managers, administrative or clerical staff, nurses, and other clinical staff or paramedics, respectively.

**Table 1.** Characteristics of the participants (n=222)

| **Variable** | **n** | **(%)** |
| --- | --- | --- |
| **Years of experience** |  |  |
| Less than 2 months | 9 | (4%) |
| 2 months to less than 1 year | 32 | (14%) |
| 1 year to less than 3 years | 47 | (21%) |
| 3 years to less than 6 years | 70 | (32%) |
| 6 years to less than 11 years | 48 | (22%) |
| 11 years or more | 16 | (7%) |
| **Staff position** |  |  |
| Physician (MD or DO) | 22 | (10%) |
| Leaders and managers | 15 | (7%) |
| Administrative or Clerical Staff | 55 | (25%) |
| Nurse (RN), Licensed Vocational Nurse (LVN), Licensed Practical Nurse (LPN) | 69 | (31%) |
| Other Clinical Staff or Clinical Support Staff | 60 | (27%) |
| Others | 1 | (<1%) |

### Single item measures and composite measures

Table 2 shows the positive perception rate of Patient Safety and Quality Issues, information exchange with other settings, and the overall positive rating of quality and patient safety compared to the AHRQ database. The results of the first two measures were superior to those of the AHRQ database, but those of the last two measures were inferior. Figure 1 shows the positive response rates for the composite measures compared to the AHRQ database and among different occupations. The positive response results of the 10 composites were lower than those in the AHRQ database.

**Figure 1:**
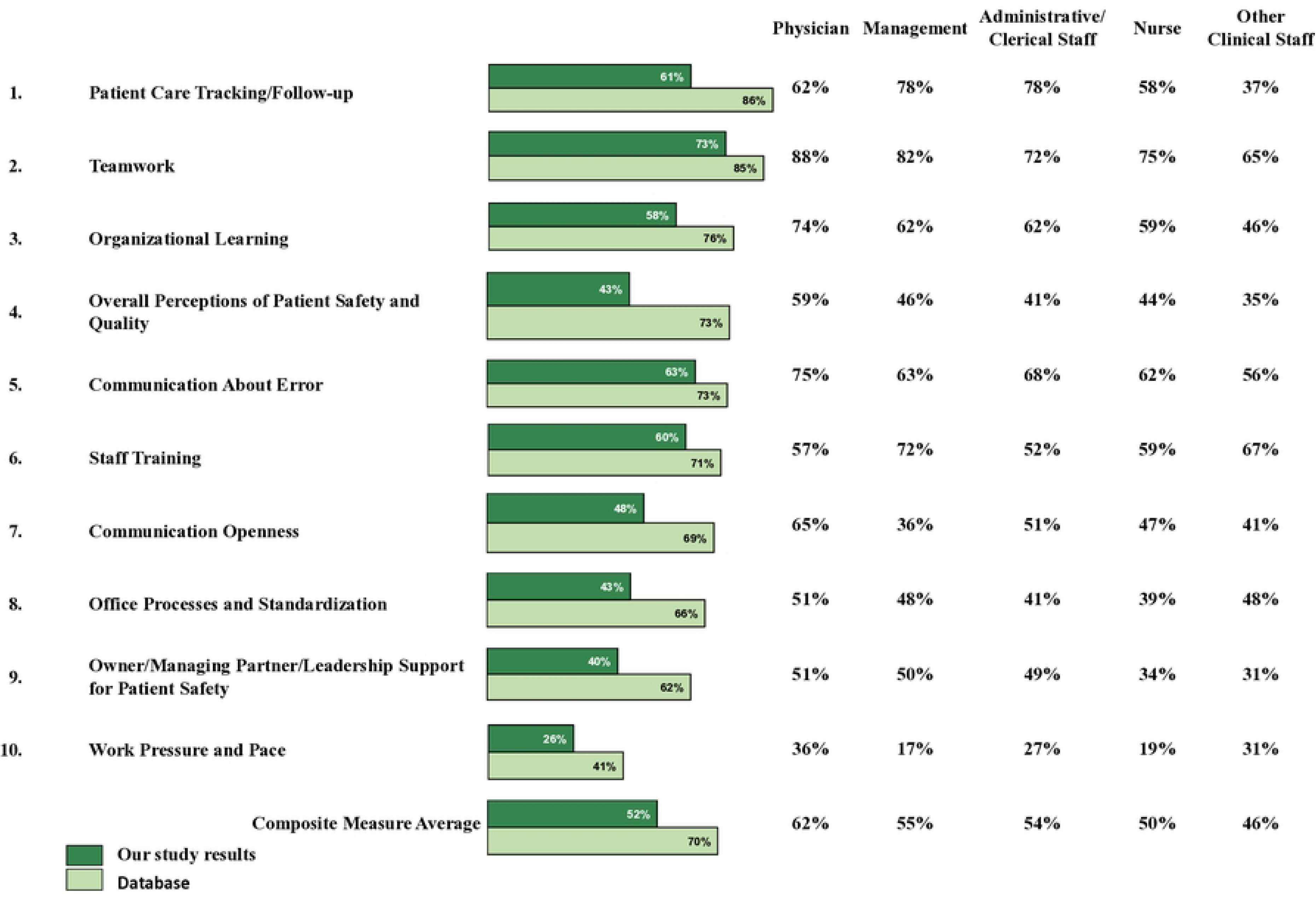
Composite measure comparative results by staff position.

**Table 2.**
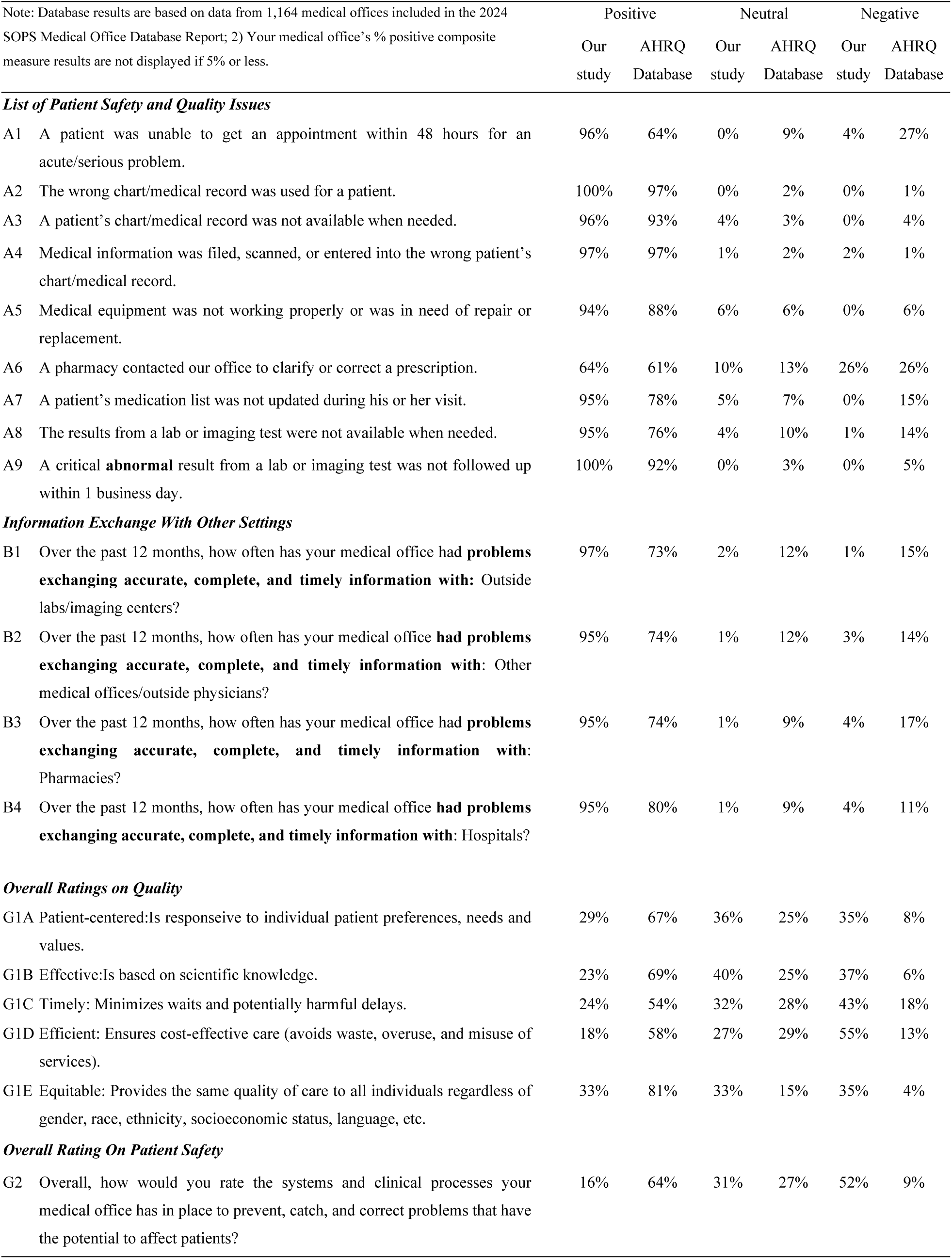
Single item measure comparative results.

### Reliability

The Cronbach’s alpha value of each composite ranged from 0.5 to 0.6 in the pilot test and above 0.6 in the actual survey, indicating better internal consistency. Additionally, the actual survey achieved good overall consistency (>0.9). AIIC ranged from 0.31 to 0.71 in the pilot test and 0.33 to 0.49 in the actual survey. Therefore, AIIC improved, indicating that the items measured the same construct. Detailed results are listed in Table 3. It also showed that our adapted version had lower Cronbach’s alpha values compared to the AHRQ reliability statistics, which is based on the pilot test data from 202 medical offices and more than 4200 staff [20]. Notably, composite 8, “Staff training,” had the lowest internal consistency (0.8 versus 0.62).

**Table 3.**
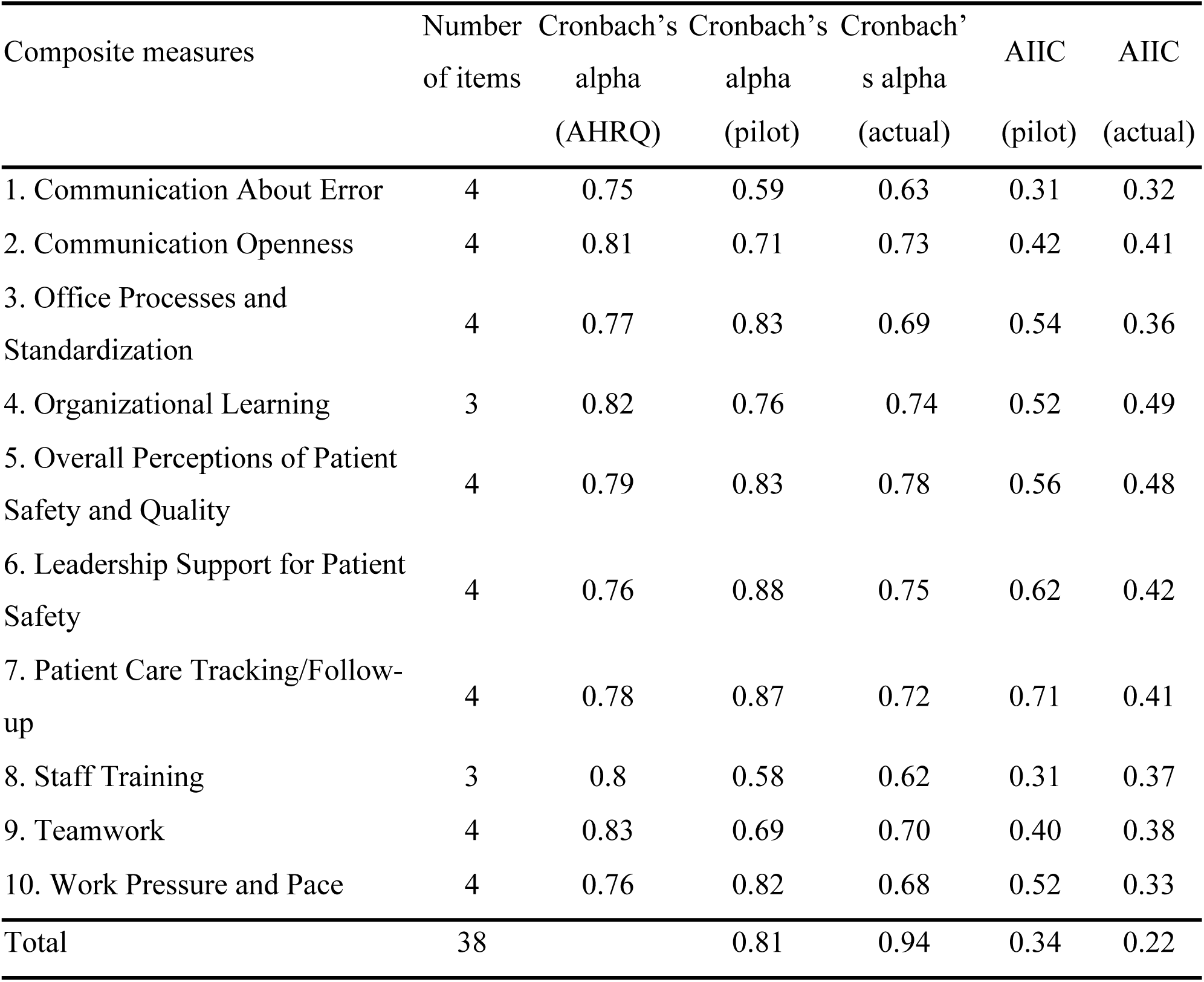
Reliability of each composite measure.

### Construct validity

A KMO of 0.74 and the significance of the Bartlett’s test of sphericity (p < 0.05) suggested that our sample size was appropriate for factor analysis. Standardized factor loadings ranged from 0.09 to 0.9, and some indicators for model fit did not meet the criteria (X^2^ = 1362.6, df = 620, p <0.001, RMSEA = 0.074 [90% confidence interval: 0.068-0.079], CFI = 0.719, SRMR = 0.108). As some factors (D7, E2, and C3) showed inadequate factor loadings (0.17, 0.09, and 0.26, respectively) and Cronbach’s alpha could be improved by dropping these items, we decided to run another CFA after deleting those items.

The new data model of 35 items was also appropriate for factor analysis (KMO=0.81, Bartlett’s test of sphericity <0.05). Standardized factor loadings ranged from 0.51 to 0.91, and goodness-of-fit indices were either excellent or acceptable (X^2^ = 1055, df = 515, p <0.001, RMSEA = 0.069 [90% confidence interval: 0.063-0.075], CFI = 0.814, SRMR = 0.087). Cronbach’s alpha values improved, but the average inter-item correlations of the composites marginally increased. Supplementary table shows the CFA results of the 38-item model and the changes in reliability statistics after three items were deleted. The deleted questions are shown in bold.

Figure 2 shows the polychoric correlations of the new 35-item model of the adapted version. Positive correlations were observed among 10 composites of patient safety culture ranging from 0.04 to 0.66. The highest correlation (r=0.66) was found between “Office Processes and Standardization” and “Work Pressure and Pace”. Conversely, the lowest correlation (r=0.05) was observed between “Organizational Learning” and “Work Pressure and Pace” as well as “Owner/Managing Partner/Leadership Support for Patient Safety” and “Teamwork”.

**Figure 2:**
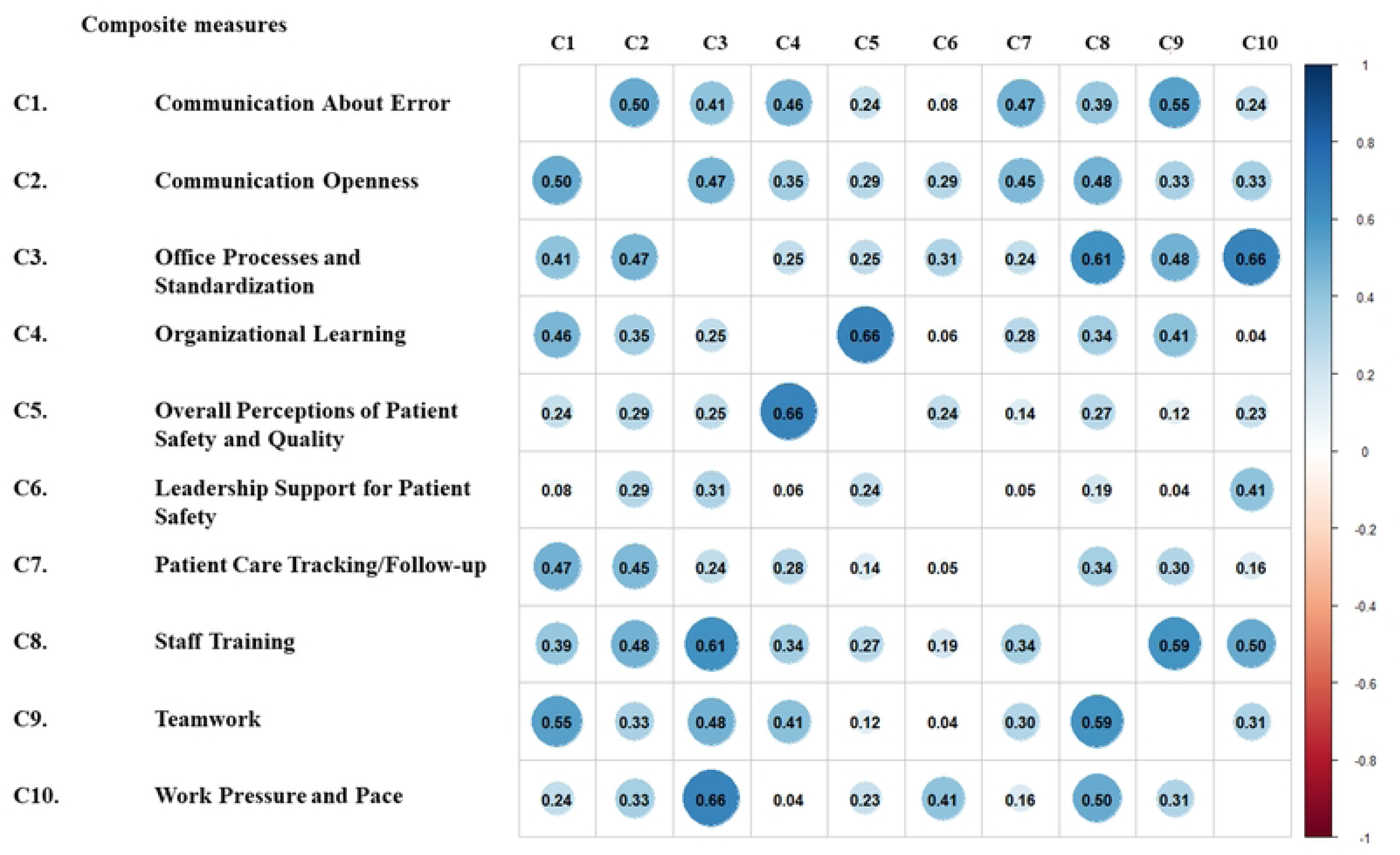
Polychoric correlations of 35-item Japanese MOSPSC.

## Discussions

This is the first study to translate, adapt, and validate the MOSPSC into Japanese. In 2011, Ito et al. culturally adapted the HSOPS for use in Japan. Their instrument had acceptable levels of internal consistency and construct validity, and the composite structures were almost identical to the original HSOPS [21]. Similarly, in our study, both the original 38-item and new 35-item models indicated good internal consistencies with a high overall Cronbach’s alpha value (>0.9), and moderate-to-high alpha values in composite measures (>0.6). AIIC values indicated that the items in the composite measured the same construct, ensuring homogeneity and unidimensionality. The model structure with 10 composites was also acceptable when evaluated in the confirmatory factor analysis.

When goodness-of-fit indices were evaluated, the 35-item model had better goodness-of-fit and internal consistency than the original 38-item model. The 35-item model was an adapted version after deleting three items with low factor loadings on composites 1, 6, and 10 from the original model. Two of the deleted items were the only negatively and positively worded items in composites 1 and 6, and the third item in composite 10 was the one we modified the sentence structure as the AHRQ pointed out that it should be tested again as the two back-translations differed. Therefore, it is necessary to investigate the wording or structure of these items in further studies on the MOSPSC in Japan.

Consistent with previous studies on cross-cultural adaptation of MOSPSC in other countries [22–25], our Japanese version of MOSPSC showed a lower internal consistency compared to the AHRQ reliability statistics, especially “Staff training”. In this composite, training on new processes, providing on-the-job training, and assigning untrained tasks were evaluated and yielded inconsistent responses, which may be responsible for the relatively low internal consistency. These results also identified areas for improvement in staff training and education.

In our Japanese version, we identified a strong patient safety dimension with regards to access to care, patient identification, availability and completeness of medical records, maintenance of medical equipment, medication updates, and timeliness of test results. Similarly, the positive rating results regarding exchanging accurate, complete, and timely information with labs, imaging centers, other medical offices, outside physicians, pharmacies, hospitals, etc. were high (>95%) and superior to the AHRQ database. However, the overall positive rating of quality was very low (18% to 33%) when compared to the AHRQ database, which ranged from 54% to 81%. The overall positive rating for patient safety was quite low, accounting for only one-fourth of the AHRQ database results (16% versus 64%). These results identified areas for improvement and maintenance of their strengths. They also indicated the need to raise awareness among leaders and staff regarding the results of the MOSPSC survey.

Among the composites of patient safety culture, teamwork was rated highest (73% positive rating), particularly among physicians (88%). “Teamwork” was also identified as the highest safety domain in a patient safety culture survey conducted using the MOSPSC in Yemen (96%), Portugal (76%), and Greece (82%) [22–24]. Conversely, work pressure and pace were poorly rated (26%), especially among leadership (17%) and nurses (19%). This might be due to an imbalance between workload and human resources, particularly among nurses. The shortage of nurses has long been an issue due to the increasing healthcare demand associated with a rapidly aging society. According to the Ministry of Health, Labor, and Welfare (MHLW), there could be a shortage of 60,000–270,000 nurses by 2025 [26]. The healthcare working environment in Japan is also extremely harsh due to free access to the healthcare system. According to 2020 OECD report, the number of outpatient visits per physician per year was 3.5 times that of the United States [27]. “Work Pressure and Pace” was also the lowest scoring dimension on patient safety culture surveys in Portugal (21%), Greece (46%), Slovenia (10.7%), and Yemen (50%) [22–25].

Our study has a few limitations. First, the survey was conducted in only four clinics in the Tokyo area. This could affect the representativeness of the sample and generalizability of the findings to other outpatient clinics. However, since it is the Japanese outpatient version of the medical safety survey questionnaire, whose reliability and validity have been verified for the first time, it will be useful for improving medical safety and quality in our medical group, as well as in other outpatient medical institutions, serving as a benchmark between facilities. Second, we did not examine other psychometric properties (e.g., convergent validity, discriminant validity, and test-retest reliability) beyond structural validity. As we plan to conduct this survey annually, we can evaluate and improve it in the future.

## CONCLUSIONS

We adapted the Japanese version of the MOSPSC and verified that the instrument had good reliability and construct validity. This is the first study to develop an instrument that can be used to measure patient safety culture, benchmark other facilities, and promote patient safety efforts in Japanese ambulatory clinics. Owing to the increasing healthcare demand, shortage of human resources, and sophistication of healthcare in Japan, our tool is very relevant for continuously monitoring safety culture and identifying areas of improvement to focus actions for improvement.

## Data Availability

All relevant data are within the manuscript and its Supporting Information files.

## Acknowledgements

The authors thank the Agency of Healthcare Research and Quality (AHRQ), Dr. Keisuke Hata, Dr. Ichiro Akagi, Dr. Ken Endo, Dr. Kazuhiro Hara, members of the expert panel, staff at four clinics in the Tokyo area: Tokyo Midtown Roppongi, Nihonbashi, Ariake and Tokyo Bay.

## 28. Supporting information

**Supplementary figure :**
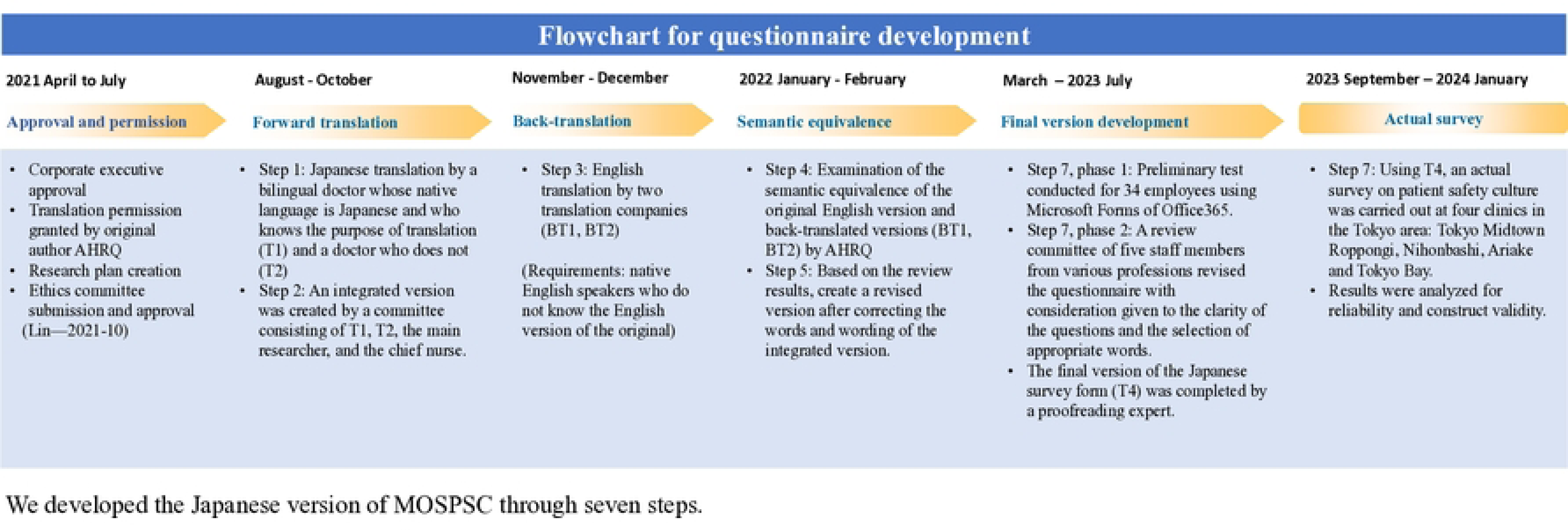
Steps involved in the development of MOSPSC Japanese version.

Supplementary data: Details of semantic equivalence assessment, pilot test and focused discussion

